# Longitudinal Humoral and Cell-Mediated Immune Responses in a Population-Based Cohort in Zurich, Switzerland between March and June 2022 - Evidence for Protection against Omicron SARS-CoV-2 Infection by Neutralizing Antibodies and Spike-specific T cell responses

**DOI:** 10.1101/2023.02.20.23286166

**Authors:** K.D. Zens, D. Llanas-Cornejo, D. Menges, J.S. Fehr, C. Münz, M.A. Puhan, A. Frei

## Abstract

**Background:** The correlate(s) of protection against SARS-CoV-2 remain incompletely defined. Additional information regarding the combinations of antibody and T cell-mediated immunity which can protect against (re)infection are needed.

**Methods:** We conducted a population-based, longitudinal cohort study including 1044 individuals of varying SARS-CoV-2 vaccination and infection statuses. We assessed Spike (S)- and Nucleocapsid (N)-IgG and wildtype, delta, and omicron neutralizing antibodies. In a subset of 328 individuals, we evaluated S, Membrane (M) and N-specific T cells. 3 months later, we reassessed antibody (n=964) and T cell (n=141) responses and evaluated factors associated with protection from (re)infection.

**Results:** At study start, >98% of participants were S-IgG seropositive. N-IgG and M/N-T cell responses increased over time, indicating viral (re)exposure, despite existing S-IgG. Compared to N-IgG, M/N-T cells were a more sensitive measure of viral exposure. N-IgG titers in the top 33% of participants, omicron neutralizing antibodies in the top 25%, and S-specific T cell responses were all associated with reduced likelihood of (re)infection over time.

**Conclusions:** Population-level SARS-CoV-2 immunity is S-IgG-dominated, but heterogenous. M/N T cell responses can distinguish previous infection from vaccination, and monitoring a combination of N-IgG, omicron neutralizing antibodies and S-T cell responses may help estimate protection against SARS-CoV-2 (re)infection.

## INTRODUCTION

It is now well-understood that exposure to SARS-CoV-2 elicits robust antibody and T cell-mediated immune responses to multiple viral proteins – in particular spike (S), nucleocapsid (N) and membrane (M) proteins^1-5^. In contrast to infection, Covid-19 vaccination elicits responses to the viral S protein; the only antigenic component of the vaccines most widely-used in the United States and Europe^6,7^. As the correlate(s) of protection, and level of said correlate(s), needed to prevent infection or severe illness have yet to be clearly defined^8^, data on population-level humoral and cellular immune responsiveness to SARS-CoV-2 remain important for understanding 1) the scope of viral exposure and 2) what proportion of the population possesses some degree of virus-specific immunity.

While much is now known regarding population-level antibody responses to SARS-CoV-2 infection, our understanding of T cell-mediated immunity is much less comprehensive. T cell responses have been described following both vaccination^9-13^ and infection; including mild or asymptomatic cases even without seroconversion^1-3,5,13-16^. However, extensive studies of T cell responses, particularly at the population level, are lacking, partially due to the labor-intensive and relatively low-throughput nature of assays designed to evaluate them, such as ELISpot and flow cytometry-based assays. To address this, adaptation of interferon (IFN)-gamma release assays (IGRAs), such as those used in *Mycobacterium tuberculosis* and Cy-tomegalovirus screening^17,18^, may aid in the detection of SARS-CoV-2-specific T cells in a larger number of samples. Importantly, as both humoral and cellular responses contribute to immunity against SARS-CoV-2, a better understanding of the heterogenous combinations of immune memory which can protect against disease may help to inform vaccination strategies, including the administration of additional booster vaccine doses.

Here, we conducted a population-based cohort study evaluating antibody and T cell responses to SARS-CoV-2 among individuals aged 16+ in Zurich, Switzerland, including individuals of varying vaccination and infection statuses. In March 2022, for all study participants (n=1044) we evaluated total SARS-CoV-2 S- and N-IgG antibody levels, as well as neutralizing antibody activity to wildtype (WT) virus and delta and omicron variants. In a randomly selected subset of individuals (n=328), we further assessed T cell responses to S, M and N proteins by IGRA. To investigate longitudinal changes in immune responses over time we reassessed antibody (n=964) and T cell (n=141) responses 3 months later, in June 2022. Overall, we found distinct immune response patterns among participants depending on reported infection and vaccination statuses. Already at the beginning of the study, nearly all participants had detectable S-IgG responses. In contrast, N-IgG and M/N-specific T cell responses increased significantly over time, in spite of existing S-IgG, indicating viral (re)exposure. Importantly, participants with N-IgG titers in the to 33%, omicron neutralizing antibody activity in the top 25%, and those with IFN-gamma-producing S-reactive T cells all had significantly reduced likelihood of (re)infection between March and June 2022. Together, our results indicate that population-level immune responses to SARS-CoV-2 are S-IgG-dominated, but heterogenous. They suggest a role for assessing M/N-specific T cells in estimating previous viral exposure, and further suggest that monitoring a combination of N-IgG, omicron neutralizing antibodies and S-reactive T cell responses may help to predict population-level protection against omicron SARS-CoV-2 (re)infection. Our findings are consistent with SARS-CoV-2-specific immunity which is mediated by co-correlates, rather than a single correlate, of protection.

## ABBREVIATED METHODS

Detailed methods and information on statistical analyses can be found in the supplementary materials.

### Participant Recruitment and Sample Collection

Individuals aged 16+ residing in the canton of Zurich, Switzerland were randomly selected by age-stratified intervals from a population registry and invited to participate. In total, 4875 individuals were contacted and 1044 enrolled (21.4% participation, Supplementary Fig.1). Initial study visits were conducted from March 1st through 31st, 2022 and second study visits (964/1044, 92.3% participation, Supplementary Fig.1) were conducted from June 7th through July 11th, 2022. At each visit, participants provided information regarding previous Covid-19 vaccination and positive SARS-CoV-2 tests. From each participant, 10mL of venous blood were collected and plasma cryopreserved prior to analysis of S-Ig and N-Ig levels and WT, delta, and omicron SARS-CoV-2 neutralizing antibody activity. For participants selected for T cell assessment, an additional 5mL of venous blood were collected and immediately used for IFN-gamma Release Assay analysis.

### Spike- and Nucleocapsid-Specific IgG and SARS-CoV-2 Neutralizing Antibody Activity

Cryopreserved plasma samples were thawed and analyzed for S- and N-specific IgG by Luminex assay as described^19^. Mean fluorescence intensity (MFI) values for each sample were divided by the mean value of negative control samples to yield an MFI ratio. Individuals were considered seropositive if the MFI ratio exceeded a lower limit of detection (LOD) of 6.0^19^. Plasma samples were further evaluated for WT, delta, and omicron SARS-CoV-2 neutralizing antibodies using a cell- and virus-free assay as described^20^. Half maximal inhibitory concentration (IC50) values of 50.0 and 2430.0 were set as lower and LODs, respectively.

### Interferon-gamma Release Assay (IGRA)

T cell responses were assessed by IGRA from whole blood stimulated overnight with overlapping 15mer peptide pools spanning the entire M and N proteins (M/N pool) or the S1 domain of the spike protein and a mix of the predicted immunodominant peptides from spike containing the majority of the S2 domain (S pool) (M, N, S1 and S PepTivator peptide pools, respectively; Miltenyi Biotec). After incubation, stimulated plasma was collected and IFN-gamma assessed using the Human IFN-gamma ELISA assay (Human IFN-gamma DuoSet ELISA kit, R&D Systems, Catalog DY285B, and DuoSet ELISA Ancillary Reagent Kit 2, R&D Systems, Catalog DY008) according to manufacturer’s instructions.

## RESULTS

### Participant Demographics and Overall Antibody and T cell Immune Responses

Of March 2022 study participants (n=1044, Supplementary Fig.1, Supplementary Table 1), 45.5% were male and 54.3% were female. 73.7% were aged 16-64 and 26.3% were 65+. 93.5% reported previous SARS-CoV-2 vaccination; 90.8% were fully vaccinated (2+ vaccine doses) and 72.1% had received at least one booster (3+ vaccine doses)^21,22^. 32.6% of participants reported a previous SARS-CoV-2 infection (defined as having received a positive PCR or antigen test result) at some point from the pandemic start up to the study visit. Older participants (65+) were more likely to report being immunized against Covid-19 (OR 2.87, 95% CI 1.36-6.09, p=0.006) and less likely to report previous infection (OR 0.44, 0.32-0.61, p<0.0001) compared to participants aged 16-64, possibly reflecting both the emphasis on vaccination for those 65+, as well as the preventative effect of vaccination on subsequent infection.

98.4% of participants were S-IgG seropositive and 23.2% were N-IgG seropositive (Fig.1A)^21,22^. 96.8%, 93.7% and 89.5% of participants had detectable neutralization IC50 values to WT, delta and omicron viral variants, respectively. Geometric mean IC50 values, however, differed significantly between variants, being highest for WT and lowest for omicron (p<0.0001 for all comparisons, Friedman test with Dunn’s multiple comparisons, Fig.1B). In a subset of study participants (n=328), circulating T cell responses to S, or a combination of M and N proteins^14^, were assessed. 89.6% had detectable S-specific T cell responses, while 57.3% had detectable M/N-specific T cell responses; geometric mean IFN-gamma production was greater for S-, as compared to M/N-stimulation (Fig.1C). Taken together, these data indicate that, as of March 2022, nearly 99% of the population had previous SARS-CoV-2 antigen exposure (either through vaccination, infection, or both). As M/N proteins are not present in available vaccines^6,7^ but are generated in response to infection, and, as M- and N-T cell responses are longer-lasting than N-IgG^1,3,14^, these findings further suggest that at least 57% of the population had been previously infected by this time.

**Figure 1.**
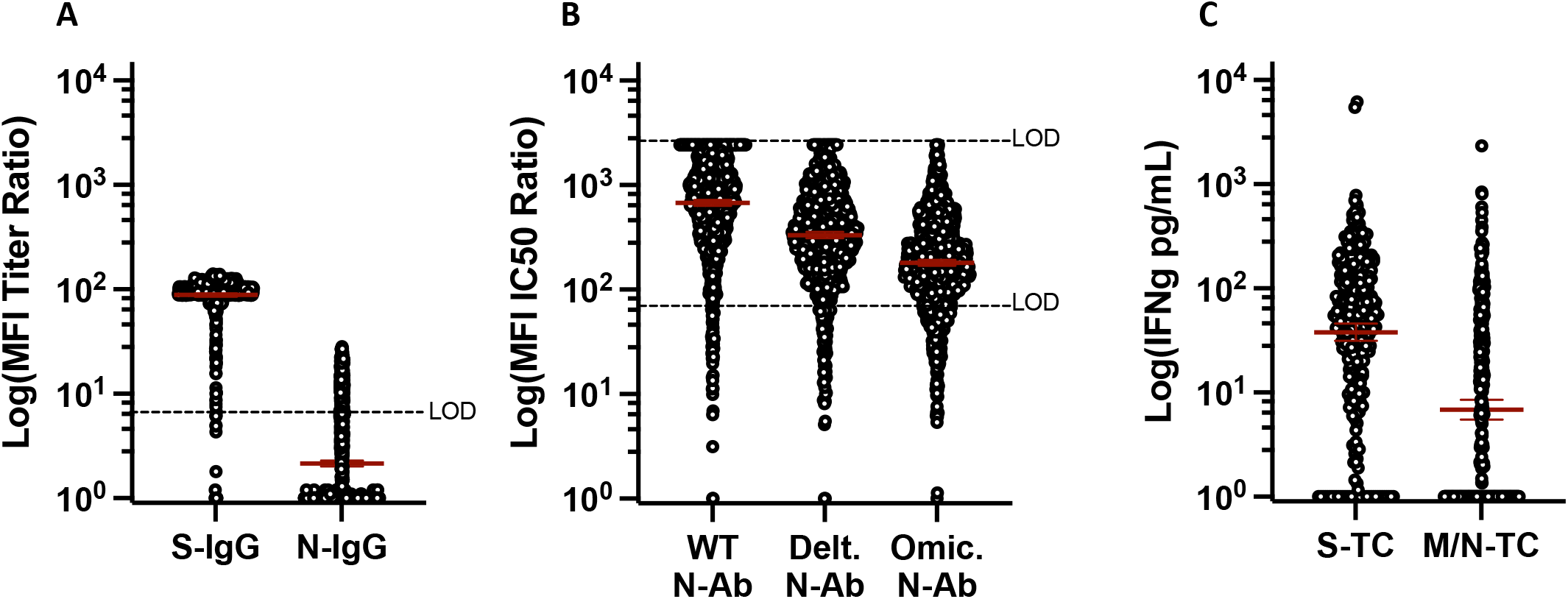
Quantitative Antibody and T cell Responses among Participants, March 2022. A) anti-S- and N-IgG geometric mean MFI titer ratios (n=1044), assay Limit of Detection (LOD)=6.0. B) anti-wildtype, delta and omicron geometric mean neutralizing antibody titers (n=1044), assay LOD 50.0-2430.0. C) Geometric mean IFN-gamma production following S or M/N peptide stimulation of whole blood (n=328).

### Impacts of Infection and Vaccination on Antibody and T cell Responses

We next assessed the impacts of infection and vaccination on antibody titers and T cell responses by multivariable linear regression. Increasing age (65+ vs. 16-64) was significantly and independently associated with lower S- and N-IgG titers, lower anti-WT, -delta, and -omicron neutralization IC50 values and lower S-T cell responses (Fig.2). Previous SARS-CoV-2 infection and receiving an increasing number of vaccine doses were both associated with significantly increased S-IgG titers, anti-WT, -delta, and -omicron neutralization IC50 values, and S-T cell responses (Fig.2). Previous infection was further associated with increased N-IgG titers and M/N-T cell responses (Fig.2). Participants were stratified into four groups: infected/vaccinated (n=285 antibody-, 80 T cell-tested), uninfected/vaccinated (n=686 antibody, 229 T cell-tested), infected/unvaccinated (n=53 antibody, 14 T cell-tested), and uninfected/unvaccinated (n=15 antibody, 3 T cell-tested) (Fig.3A and B). S-IgG and neutralizing activity tended to be higher in vaccinated individuals (both previously infected and uninfected, Fig.3A), and these did not correlate with N-IgG responses (Fig.3B). In contrast, N-IgG and M/N-T cell responses were higher among previously infected individuals (both vaccinated and unvaccinated, Fig.3A and B). Unsurprisingly, the lowest overall responses were observed in uninfected/unvaccinated individuals and, due to low sample number, there were insufficient data to assess T cell correlation patterns for this group (Fig.3A and B). In general, however, antibody and T cell response patterns appeared more similar between vaccinated participants compared to infected participants, with robust S-IgG and neutralizing antibody responses following vaccination and greater N-IgG and M/N-T cell responses among infected individuals.

**Figure 2.**
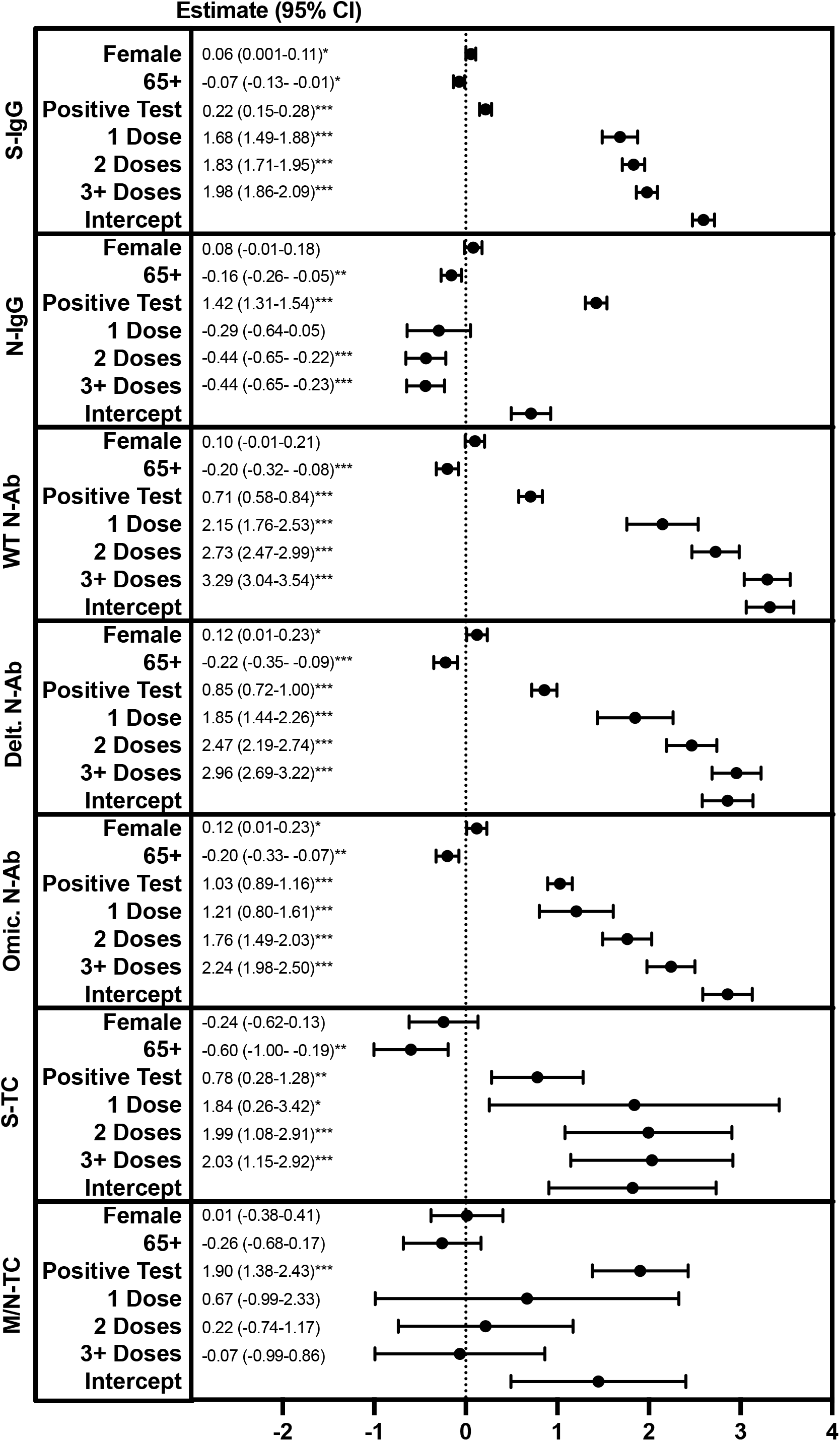
Factors Associated with March 2022 S- or N-IgG, N-Ab Titers, or S- or M/N-T cell IFN-gamma Levels. Multivariable linear regression modeling was used to assess the relationship between gender (female vs. male), age group (65+ vs. 16-64 years), reporting a previous SARS-CoV-2 infection (positive PCR or antigen test) (yes vs. no), and the number of Covid-19 vaccine doses received (1, 2, 3+ vs. 0), and S- or N-IgG, N-Ab MFI ratio titers, or S- or M/N-T cell IFN-gamma levels (natural logarithm-transformed). *p>0.05, **p>0.01, ***p>0.005.

**Figure 3.**
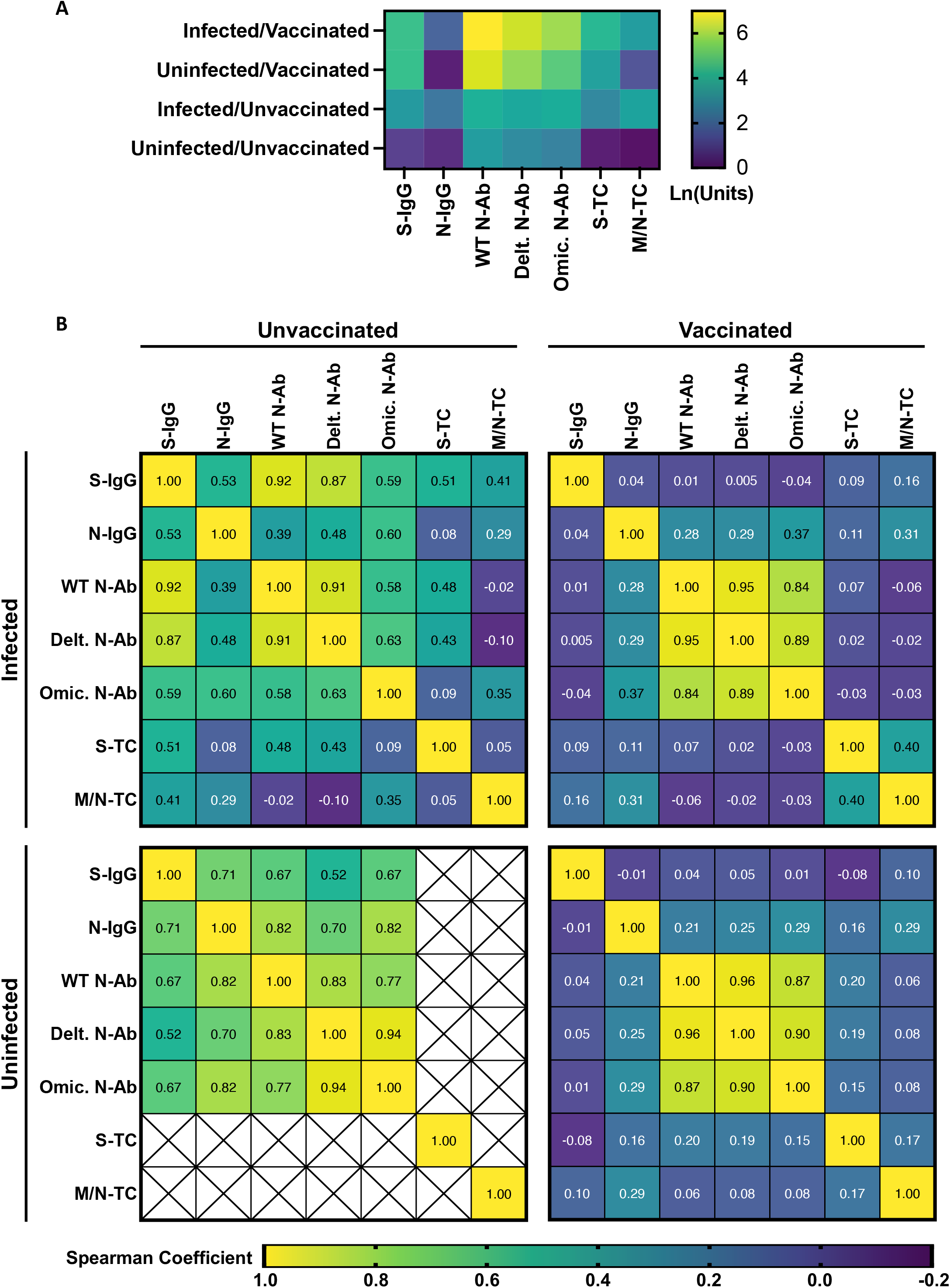
Antibody and T cell Responses among Participants by Infection and Vaccination Status, March 2022. A) Quantitative Antibody and T cell Responses. Log10 anti-S- and N-IgG geometric mean MFI titer ratios, anti-wildtype, delta and omicron geometric mean neutralizing antibody titers and geometric mean IFN-gamma production following S or M/N peptide stimulation of whole blood. B) Correlation between Antibody and T cell Responses. Top left: Infected and Unvaccinated individuals (n=53 antibody tested, 14 T cell tested), Top right: Infected and Vaccinated (n=285 antibody tested, 80 T cell tested), Bottom left: Uninfected and Unvaccinated (n=15 antibody tested, 3 T cell tested), Bottom right: Uninfected and Vaccinated (n=686 antibody tested, 229 T cell tested). Values represent Spearman correlation coefficients for indicated antibody and T cell response pairs. Crosses indicate pairs with insufficient data for analysis.

### Longitudinal Responses and Protection from (Re)infection

964 participants returned for a second study visit, 3 months later, in June 2022 (Supplementary Fig.1, Supplementary Table 1). 141 were assessed for T cell responses (118 longitudinally from March and an additional 23 not evaluated for T cell responses in the March round; Supplementary Table 1). At this time, 6.4% of participants were unvaccinated, 2.4% had received a single vaccine dose, 17.0% had received 2 doses, and 74.2% had received 3 or more doses. 19 individuals (2.0% of the study population) received an additional vaccination between March and June, all of which were 2nd or booster doses. 16.0% reported a positive SARS-CoV-2 PCR or antigen test (infection) between March and June. Of these, 14.3% also reported previous infection in March (repeated infections; 85.7% new infections). In total, 45.3% of the population reported at least one SARS-CoV-2 infection.

98.8% of participants were S-IgG seropositive (similar to March) and 36.7% were N-IgG seropositive (increasing from March; p<0.0001, Two-sample test of proportions). Geometric mean MFI ratio titers for both S-IgG and N-IgG increased between March and June (Fig.4A; S-IgG p<0.0001, N-IgG p<0.0001, Wilcoxon matched-pairs signed rank test). 97.2% and 72.3% of participants had detectable S- and M/N-T cell responses, respectively; significantly more than in March (Fig.4A and B, S p=0.043, M/N p=0.001, Fisher’s exact test) and geometric mean IFN-gamma production among the overall population was higher for both in June (Fig.4B, S p=0.05, M/N p=0.053, Mann-Whitney test). M/N-T cell responses tended also to be higher in the longitudinal subset, though this was not statistically significant (p=0.109, Wilcoxon matched-pairs signed rank test). Between March and June, 0.4% (4/948) of those who were seropositive for S-IgG became seronegative while 50% (8/16) of those who were seronegative became seropositive. For N-IgG, 32.3% (72/223) of those who were seropositive became seronegative and 27.4% (203/741) of those who were seronegative became seropositive. Of individuals tested longitudinally for T cell responses, 1.9% (2/107) of those S-T cell positive in March were negative in June, while 81.8% (9/11) of those negative in March were positive in June. 13.3% (8/60) of those who were M/N-T cell positive in March were negative in June, while 58.6% (34/58) of those negative in March were positive in June. Of those that became N-IgG seropositive or M/N-T cell reactive, only 59.1% (120/203) and 29.4% (10/34), respectively, reported infection, highlighting the importance of immune monitoring efforts in understanding SARS-CoV-2 exposures.

**Figure 4.**
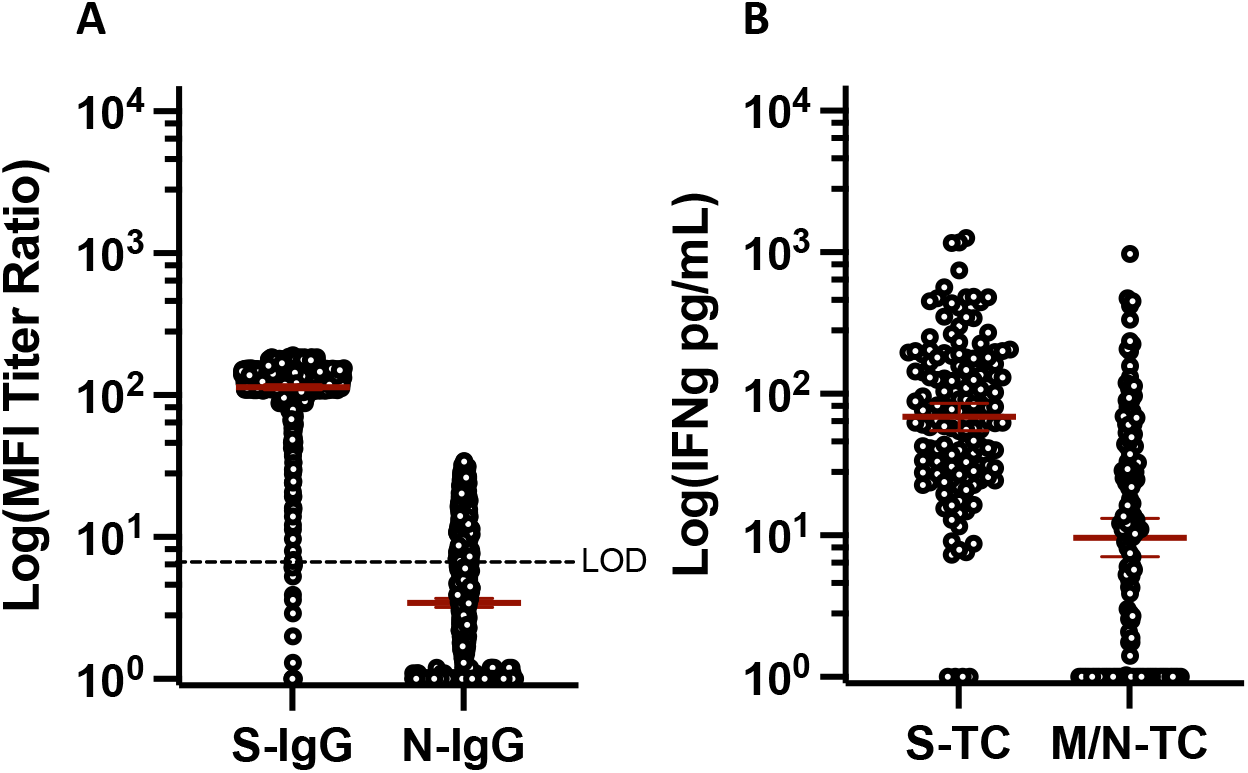
Quantitative Antibody and T cell Responses, June 2022. A) S- and N-IgG geometric mean MFI ratio titers (n=964). B) Geometric mean IFN-gamma production following S or M/N peptide stimulation of whole blood (n=141).

Overall, we observed three dominant immune response patterns among participants, which were similar in both March and June. In March we observed 17.7% group1 (S-IgG+/N-IgG+/S-T cell+/M/N-T cell+ -> positive for all factors), 36.3% group2 (S-IgG+/N-IgG-/S-T cell+/M/N-T cell+ -> positive for everything except N-IgG), and 34.2% group3 (S-IgG+/N-IgG-/S-T cell+/M/N-T cell--> S-IgG and S-T cell positive only). In June, we observed 30.5% group1, 39.0% group2, and 23.4% group3 (Supplementary Fig.2A and B). Interestingly, only 5.2% of those in group1 (positive of all factors) reported an infection between March and June compared to 26.6% of those in group2 (positive for everything except N-IgG) and 21.9% of those in group3 (S-IgG and S-T cell positive only), potentially suggesting superior protection by the group1 combination of immune responses. In order to evaluate which immune response components might be capable of providing protection against (re)infection, we assessed whether an individual’s levels of S- and N-IgG and S- and M/N-T cells in March were associated with infection between March and June (Fig.5). As vaccination is expected to influence SARS-CoV- 2-specific immune responses, individuals vaccinated between March and June (n=19) were excluded from the analysis. Having an N-IgG MFI ratio titer above 10 (titers in the top 33% of the population) was associated with an 84% reduced odds of infection between March and June (OR 0.16, 95% 0.03-0.85, p=0.031; compared to the lowest 33%). Having omicron neutralizing IC50 titers above 360 (titers in the top 25% of the population) was associated with a 94% reduced odds of infection (OR 0.06, 0.006-0.60, p=0.017; compared to the lowest 25%), while having S-T cells was associated with a 60% reduced likelihood of infection (production of >=25 to <65pg/mL IFN-gamma, OR 0.39, 0.17-0.92, p=0.030; compared to the lowest 25%). Therefore, N-IgG, omicron neutralizing antibodies, and S-specific T cells are associated with protection from omicron (re)infection, and monitoring a combination of these responses may aid in the assessment of population-level immunity against omicron SARS-CoV-2 (re)infection.

**Figure 5.**
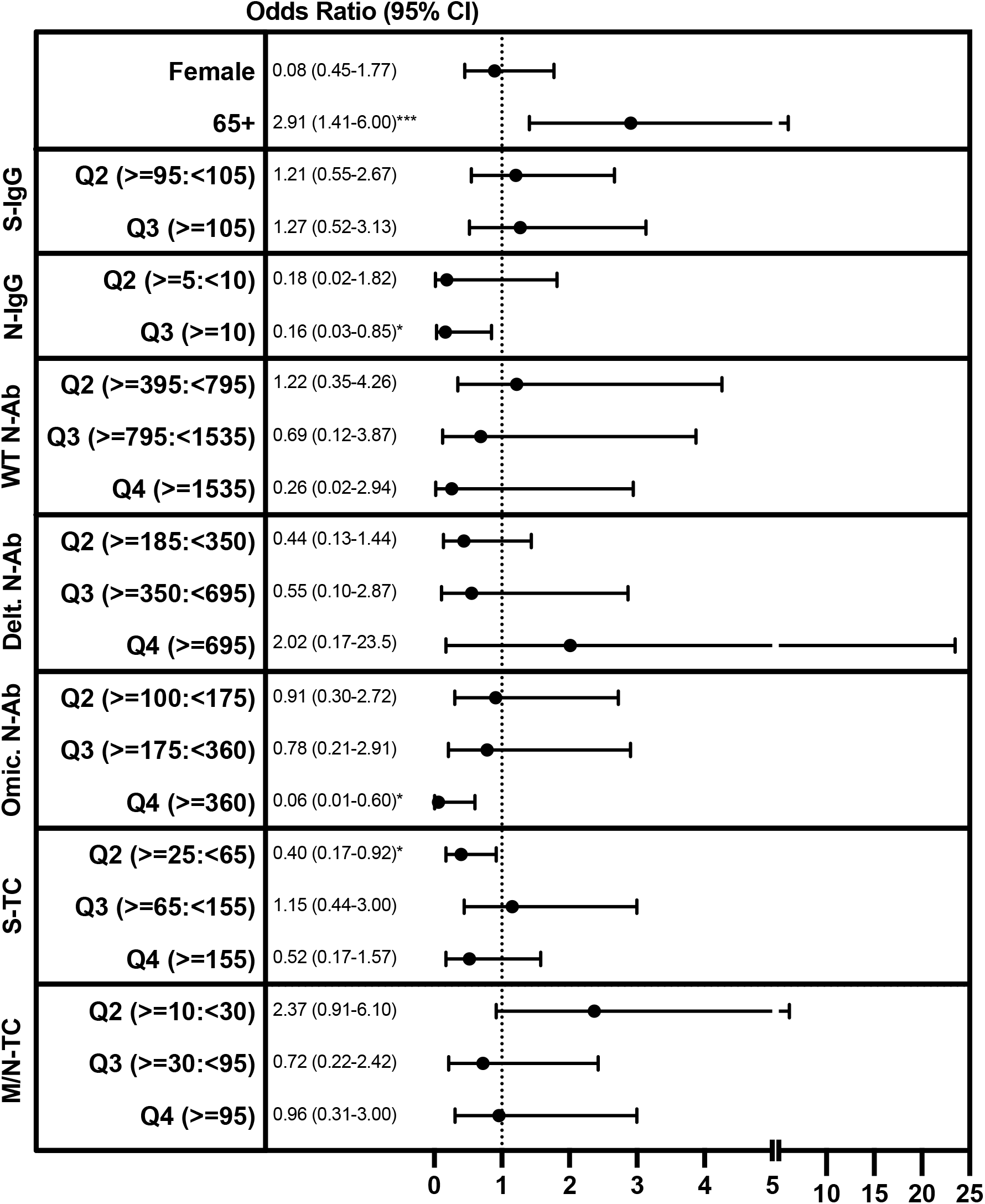
Factors Associated with Reporting a SARS-CoV-2 Infection between March and June 2022. Multivariable logistic regression modeling was used to assess the relationship between gender (female vs. male), age group (65+ vs. 16-64 years), quantiles of S- or N-IgG, N-Ab Titers, or S- or M/N-T cell IFN-gamma levels in March 2022, and reporting a previous SARS-CoV-2 infection (positive PCR or antigen test) (yes vs. no) in June/July 2022. For S- and N-IgG MFI ratio titers, individuals were assigned to one of three expression quantiles (<33%, 33-67%, 67+% of all participants); for N-Ab IC50 values, and S- and M/N-T cell IFN-gamma levels, individuals were assigned to one of four expression quantiles (<25%, 25->50%, 50->75%, 75+% of all participants). Corresponding MFI ratios/IC50 titers/IFN-gamma levels are listed next to each variable. *p>0.05, ***p>0.005.

## DISCUSSION

While antibody responses among individuals in the Zurich area, and throughout Switzerland have been well-described^21-24^, much less is known regarding population-level T cell responsiveness to SARS-CoV-2. Here, we utilized an interferon (IFN)-gamma release assay (IGRA) based on short-term culture of whole blood with SARS-CoV-2-specific peptides to assess T cell responses, which demonstrated good concordance with an ELISpot assay which we used previously^14^. We found that, by June 2022, 97% and 72%, of study participants had S- and M/N-specific T cells, respectively. In comparison, 99% of participants were S-IgG seropositive and slightly less than 40% were N-IgG seropositive. That S-specific antibody and T cell responses were higher, in general, than N-specific antibody and M/N-specific T cell responses is consistent with the high vaccination coverage in the population (>90% fully vaccinated), as available vaccines contain S, but not M or N antigens^6,7^. Furthermore, the half-life of S-IgG is substantially longer than that of N-IgG^14,25,26^, consistent with our observation that the fraction of participants who were initially seropositive in March but became seronegative by June was greater for N-IgG (14.1%) compared to S-IgG (1.2%). The higher percentage of M/N-T cell positivity compared to N-IgG positivity is worth noting and is possibly due to 1) the use of both M- and N-specific peptides in the IGRA, 2) that the half-lives of circulating M- and N-specific T cells are longer than that of N-IgG^1,3,14^, 3) that some individuals develop only M- and N-specific T cell responses after infection^14^, and, 4) that previous exposure to endemic human coronaviruses (HCoV)-229E, -NL63, -OC43, and -HKU1) can generate low levels of T cells cross-reactive to SARS-CoV-2^2,4,5^. Our findings, however, indicate that assessing SARS-CoV-2 M/N-T cells is likely a more sensitive method for evaluating exposure to SARS-CoV-2 compared to N-IgG and that monitoring M/N-T cells may help to assess population-level “hybrid immunity”.

An additional takeaway from our findings is that the majority of individuals had more than one type of virus-specific memory response and that protection was not clearly mediated only by a single subset. The most common patterns, representing over 90% of the study population were group1 (S-IgG+/N-IgG+/S-T cell+/M/N-T cell+), group2 (S-IgG+/N-IgG+/S-T cell+/M/N-T cell-), and group3 (S-IgG+/N-IgG-/S-T cell+/M/N-T cell-). All of these patterns included S-IgG, but nearly 95% of those in group1 did not report an infection between March and June compared to 73-78% of those in groups 2 and 3. Furthermore, despite high S-IgG seropositivity already in March 2022, the percentage of participants with detectable N-IgG titers and M/N-T cells increased significantly by June 2022, indicating continued viral (re)infections, even among individuals with some level of S-specific immunity. In assessing potential mediators of protective immunity, we found that having a high N-IgG titer and/or high omicron neutralizing antibody activity were both protective against omicron SARS-CoV-2 infection. As the half-life of N-IgG is relatively short at approximately 60-90 days^14,25,26^, individuals with high titers were likely recently infected – perhaps in the 3-6 months prior to March. It would also make sense that these individuals were infected with the omicron variant, which was responsible for >99% of reported Covid-19 cases in late January 2022 in Switzerland^27^. As recent infection may contribute to a state of “trained immunity”^28^ with enhanced baseline activation of the innate immune system, we speculate that N-IgG is likely not a sole mediator of protection in and of itself, but may serve as a marker for a persisting “antiviral” state which, in turn, limits reinfection. We additionally found that S-T cells were associated with reduced likelihood of infection. S-reactive T cells are known to be generated following SARS-CoV-2 infection and SARS-CoV-2 vaccination^1-3,9-15^, and individuals can also possess pre-existing memory T cell responses generated from previous endemic human coronavirus exposure^2,4,5^. Though a role for S-T cell responses as a correlate of protection against SARS-CoV-2 is not completely clear^8^, it is known that T cell-mediated immunity is more cross-reactive than corresponding antibody responses^29^. Furthermore, it has been observed in animal models that, in the absence of antibody responses, protection from SARS-CoV-2 can be mediated solely by T cell immunity^30^, and, similarly, we recently observed that individuals can clear SARS-CoV-2 infection in the absence of detectable antibody responses^14^, highlighting the importance of this subset in protection from infection.

Some limitations to our study include, first, that we relied on self-reported SARS-CoV-2 infections based on receiving a positive PCR or antigen test result. While false positive results are possible, it is also likely that, as many individuals use self-tests, which have limited sensitivity especially early in infection, that true infections are under-reported. Similarly, we observed that a substantial fraction - 20% (3/15) - of participants that reported being uninfected/unvaccinated had detectable S- or N-IgG titers, which we would expect only in responses to SARS- CoV-2 antigen exposure. An additional limitation was the low number of uninfected/unvaccinated individuals, and as we collected T cell data only from a subset of individuals, we did not have sufficient data to thoroughly assess this group, which represents an interesting immune “baseline”. Furthermore, in terms of the assays used, we assessed only IFN-gamma as a measure of T cell activity. Studies have demonstrated that IL-2-producing T cell responses are also generated in SARS-CoV-2 infection and vaccination^5,16^. It would be valuable to test the IGRA approach in evaluating IL-2 responses in further studies as well. In addition, we limited our T cell analysis to the three dominant antigens for cellular immune responses (S, M and N), and furthermore, for experimental feasibility, S1 and S2 domains as well as M and N responses were pooled, although they have been shown in other studies to exhibit some distinct behaviors^1,2,14,16^. We cannot exclude the importance of subdominant T cell responses against other viral antigens in some of the participants, which may have led to an underestimation of T cell responses. Another limitation is that, although the IGRA results had a high degree of concordance with ELISpot assay (Supplementary Methods, Supplementary Fig3A and B), they did not strongly correlate. This is not unexpected, though, as IGRA measures total IFN-gamma output in pg/mL which could be produced by few specific T cells, while ELISpot assesses only the number of IFN-gamma-producing cells without taking the amount of IFN-gamma produced by individual cells into account, making it difficult to compare values from these two assays directly. Furthermore, we used a surrogate assay to indirectly quantify neutralizing activity by measuring competitive inhibition of trimeric SARS-CoV-2 S protein binding to the Angiotensin Converting Enzyme 2 (ACE2) receptor. However, this assay showed high sensitivity compared to live virus assays during validation^20^ and permitted simultaneous assessment of neutralization against WT-SARS-CoV-2, delta and omicron variants.

Nevertheless, we provide here population-level estimates of cellular immunity as well as factors which may be associated with protection from omicron SARS-CoV-2 infection. Our results suggest that, while the majority of individuals possess anti-S-IgG, these responses in and of themselves are likely not a good predictor of protection from (re)infection. In terms of estimating what fraction of the population has been infected with SARS-CoV-2, monitoring M/N-reactive T cells responses may be helpful. However, to assess what fraction of the population might be protected from (re)infection with omicron SARS-CoV-2, our data suggest that monitoring a combination of anti-N-IgG responses, omicron-specific neutralizing antibody responses, and S-reactive T cell responses may be beneficial. Our findings indicate a pattern where co-correlates of protection, rather than simply S-IgG, are likely important for mediating long-term protective immunity against SARS-CoV-2 and future variants and provide important information for policy makers regarding vaccination strategy in the case of changing disease epidemiology.

## Data Availability

All data produced in the present study are available upon reasonable request to the authors and will be made available upon study publication.

## CONFLICT OF INTEREST

The authors declare no competing interests.

## FUNDING

The Corona Immunitas research network is coordinated by the Swiss School of Public Health (SSPH+) and funded by fundraising of SSPH+ including funds of the Swiss Federal Office of Public Health and private funders (ethical guidelines for funding stated by SSPH+ were respected), by funds of the cantons of Switzerland (Vaud, Zurich, and Basel), and by institutional funds of the Universities.

## ETHICAL APPROVAL

We obtained written, informed consent from all participants upon study enrollment. Participants were compensated with a flat fee for any travel expenses related to study visits, but otherwise did not receive any compensation for their participation. The study protocol was approved by the Cantonal Ethics Committee of Zurich (BASEC Registration No. 2020-01247) and registered (ISRCTN registry 18181860, date of registration 13 July 2020, retrospectively registered).

## ACKNOWLEDGEMENTS

The authors would like to thank Danusia Vanoaica and Osman Yoztekin for their support with the T cell assays, and the study administration team and the study participants for their dedicated contribution to this research project.

## Notes

### Competing Interest Statement

The authors have declared no competing interest.

### Author Declarations

The study protocol was approved by the Cantonal Ethics Committee of Zurich (BASEC Registration No. 2020-01247) and registered (ISRCTN registry 18181860, date of registration 13 July 2020, retrospectively registered).

## REFERENCES

1 Dan, J. M. et al. Immunological memory to SARS-CoV-2 assessed for up to 8 months after infection. Science 371, eabf4063 (2021). https://doi.org:doi:10.1126/science.abf4063

2 Grifoni, A. et al. Targets of T Cell Responses to SARS-CoV-2 Coronavirus in Humans with COVID-19 Disease and Unexposed Individuals. Cell 181, 1489–1501.e1415 (2020). https://doi.org:https://doi.org/10.1016/j.cell.2020.05.015

3 Cohen, K. W. et al. Longitudinal analysis shows durable and broad immune memory after SARS-CoV-2 infection with persisting antibody responses and memory B and T cells. Cell Reports Medicine 2, 100354 (2021). https://doi.org:https://doi.org/10.1016/j.xcrm.2021.100354

4 Le Bert, N. et al. SARS-CoV-2-specific T cell immunity in cases of COVID-19 and SARS, and uninfected controls. Nature 584, 457–462 (2020). https://doi.org:10.1038/s41586-020-2550-z

5 Sekine, T. et al. Robust T Cell Immunity in Convalescent Individuals with Asymptomatic or Mild COVID-19. Cell 183, 158–168.e114 (2020). https://doi.org:10.1016/j.cell.2020.08.017

6 Creech, C. B., Walker, S. C. & Samuels, R. J. SARS-CoV-2 Vaccines. JAMA 325, 1318–1320 (2021). https://doi.org:10.1001/jama.2021.3199

7 Amanat, F. & Krammer, F. SARS-CoV-2 Vaccines: Status Report. Immunity 52, 583–589 (2020). https://doi.org:https://doi.org/10.1016/j.immuni.2020.03.007

8 Goldblatt, D., Alter, G., Crotty, S. & Plotkin, S. A. Correlates of protection against SARS-CoV-2 infection and COVID-19 disease. Immunol Rev 310, 6–26 (2022). https://doi.org:10.1111/imr.13091

9 Goel, R. R. et al. mRNA vaccines induce durable immune memory to SARS-CoV-2 and variants of concern. Science 374, abm0829 (2021). https://doi.org:doi:10.1126/science.abm0829

10 Zhang, Z. et al. Humoral and cellular immune memory to four COVID-19 vaccines. Cell 185, 2434–2451.e2417 (2022). https://doi.org:https://doi.org/10.1016/j.cell.2022.05.022

11 Mateus, J. et al. Low-dose mRNA-1273 COVID-19 vaccine generates durable memory enhanced by cross-reactive T cells. Science 374, eabj9853 (2021). https://doi.org:doi:10.1126/science.abj9853

12 Tarke, A. et al. SARS-CoV-2 vaccination induces immunological T cell memory able to cross-recognize variants from Alpha to Omicron. Cell 185, 847–859.e811 (2022). https://doi.org:https://doi.org/10.1016/j.cell.2022.01.015

13 Sette, A. & Crotty, S. Immunological memory to SARS-CoV-2 infection and COVID-19 vaccines. Immunological Reviews 310, 27–46 (2022). https://doi.org:https://doi.org/10.1111/imr.13089

14 Menges, D. et al. Heterogenous humoral and cellular immune responses with distinct trajectories post-SARS-CoV-2 infection in a population-based cohort. Nature Communications 13, 4855 (2022). https://doi.org:10.1038/s41467-022-32573-w

15 Rodda, L. B. et al. Functional SARS-CoV-2-Specific Immune Memory Persists after Mild COVID-19. Cell 184, 169–183.e117 (2021). https://doi.org:https://doi.org/10.1016/j.cell.2020.11.029

16 Thieme, C. J. et al. Robust T Cell Response Toward Spike, Membrane, and Nucleocapsid SARS-CoV-2 Proteins Is Not Associated with Recovery in Critical COVID-19 Patients. Cell Reports Medicine 1, 100092 (2020). https://doi.org:https://doi.org/10.1016/j.xcrm.2020.100092

17 Pai, M. et al. Gamma Interferon Release Assays for Detection of Mycobacterium tuberculosis Infection. Clinical Microbiology Reviews 27, 3–20 (2014). https://doi.org:doi:10.1128/CMR.00034-13

18 Giulieri, S. & Manuel, O. QuantiFERON®-CMV assay for the assessment of cytomegalovirus cell-mediated immunity. Expert Review of Molecular Diagnostics 11, 17–25 (2011). https://doi.org:10.1586/erm.10.109

19 Fenwick, C. et al. Changes in SARS-CoV-2 Spike versus Nucleoprotein Antibody Responses Impact the Estimates of Infections in Population-Based Seroprevalence Studies. J Virol 95 (2021). https://doi.org:10.1128/jvi.01828-20

20 Fenwick, C. et al. A high-throughput cell- and virus-free assay shows reduced neutralization of SARS-CoV-2 variants by COVID-19 convalescent plasma. Science Translational Medicine 13, eabi8452 (2021). https://doi.org:doi:10.1126/scitranslmed.abi8452

21 Amati, R. et al. Functional immunity against SARS-CoV-2 in the general population after a booster campaign and the Delta and Omicron waves, Switzerland, March 2022. Euro Surveill 27 (2022). <https://doi.org:10.2807/1560-7917.Es.2022.27.31.2200561>

22 Frei, A. et al. Development of hybrid immunity during a period of high incidence of infections with Omicron subvariants: A prospective population based multi-region cohort study. medRxiv, 2022.2010.2014.22281076 (2022). https://doi.org:10.1101/2022.10.14.22281076

23 West, E. A. et al. Seroprevalence of SARS-CoV-2 antibodies, associated factors, experiences and attitudes of nursing home and home healthcare employees in Switzerland. BMC Infectious Diseases 22, 259 (2022). https://doi.org:10.1186/s12879-022-07222-8

24 Stringhini, S. et al. Seroprevalence of anti-SARS-CoV-2 antibodies 6 months into the vaccination campaign in Geneva, Switzerland, 1 June to 7 July 2021. Euro Surveill 26 (2021). https://doi.org:10.2807/1560-7917.Es.2021.26.43.2100830

25 Lumley, S. F. et al. The Duration, Dynamics, and Determinants of Severe Acute Respiratory Syndrome Coronavirus 2 (SARS-CoV-2) Antibody Responses in Individual Healthcare Workers. Clin Infect Dis 73, e699–e709 (2021). https://doi.org:10.1093/cid/ciab004

26 Wheatley, A. K. et al. Evolution of immune responses to SARS-CoV-2 in mild-moderate COVID-19. Nature Communications 12, 1162 (2021). https://doi.org:10.1038/s41467-021-21444-5

27 COVID-19 Switzerland Information on the current situation, Epidemiological course, Switzerland and Liechtenstein, <https://www.covid19.admin.ch/en/epidemiologic/virus-variants> (2022).

28 Netea, M. G. et al. Defining trained immunity and its role in health and disease. Nature Reviews Immunology 20, 375–388 (2020). https://doi.org:10.1038/s41577-020-0285-6

29 Sewell, A. K. Why must T cells be cross-reactive? Nature Reviews Immunology 12, 669–677 (2012). https://doi.org:10.1038/nri3279

30 Kingstad-Bakke, B. et al. Vaccine-induced systemic and mucosal T cell immunity to SARS-CoV-2 viral variants. Proceedings of the National Academy of Sciences 119, e2118312119 (2022). https://doi.org:doi:10.1073/pnas.2118312119

