## Supplementary Methods for "Longitudinal Humoral and Cell-Mediated Immune Responses in a Population-Based Cohort in Zurich, Switzerland between March and June 2022 - Evidence for Protection against Omicron SARS-CoV-2 Infection by Neutralizing Antibodies and Spike-specific T cell responses"

Participant Selection and Recruitment

For this prospective, population-based cohort study, individuals aged 16+ residing in the Canton of Zurich, Switzerland were randomly selected. The Swiss Federal Office of Statistics, which maintains address information for all registered residents of the country of Switzerland, provided random samples of the general population in age-stratified (16–29, 30–44, 45–64, and ≥ 65 years) groups. Given a sensitivity of 97% and specificity of 99% for the Luminex-based antibody used[19], we deemed 200 participants for each stratum to provide precise enough estimates for an expected seroprevalence of 90% or more, resulting in a target sample size of 800 individuals. Selected individuals were invited to in-person study visits at a healthcare facility to provide a blood sample. Individuals not willing or able to travel were offered at-home visits. For each participant, trained personnel collected venous blood samples, according to clinical standards and Covid-19 hygiene measures. Before the first study visit, all participants completed a baseline questionnaire. In total, 4875 individuals were contacted and 1044 participants were enrolled (participation rate: 21.4%). Participant enrollment and subsequent study visits were conducted between 1 and 31 March 2022. All study participants were invited for a second study visit to provide a blood sample between 7 June and 11 July 2022, of which 92.3% (964/1044) participated.

Ethical Considerations

We obtained written, informed consent from all participants upon study enrollment. Participants were compensated with a flat fee for any travel expenses related to study visits, but otherwise did not receive any compensation for their participation. The study protocol was approved by the Cantonal Ethics Committee of Zurich (BASEC Registration No. 2020-01247) and registered (ISRCTN registry 18181860, date of registration 13 July 2020, retrospectively registered).

Questionnaire Data

Participants provided information regarding previous SARS-CoV-2 vaccination and previous positive Covid-19 tests through electronic questionnaires. We used the Research Electronic Data Capture (REDCap) platform for data collection and management[31,32].

Blood Collection and Plasma and PBMC Isolation

From each participant, 10mL of venous blood were collected into a single K2-EDTA vacutainer tube (BD). From participants selected for analysis of T cell responses, an additional 5mL of venous blood were collected into a Li-Heparin vacutainer tube, respectively (BD). For K2-EDTA tubes, plasma was collected by initial, low-speed centrifugation, followed by isolation of peripheral blood monocytic cells (PBMCs) from the remaining cellular fraction by density-gradient centrifugation using Ficoll–Paque (density 1.077g/mL). Plasma aliquots were stored at -20°C prior to antibody analyses. PBMCs were initially frozen in 90% fetal bovine serum (FBS, Pan Biotech) with 10% dimethyl sulfoxide (DMSO, Sigma) at -80°C and transferred to -150°C for storage prior to use in ELISpot assays. Whole blood from Li-Heparin tubes were aliquoted and immediately used for IFN-gamma Release Assay analysis (below).

Analysis of Spike- and Nucleocapsid-Specific IgG

Cryopreserved plasma samples were thawed and analyzed for levels of Spike (S)-specific, or Nucleocapsid (N)-specific IgG by Luminex assay[19]. Assay beads were prepared by covalent coupling of either the SARS-CoV-2 Spike protein trimer, or Nucleocapsid protein, with Mag-Plex beads using a Bio-Plex 356 Amine Coupling Kit (Bio-Rad, Catalog 10000148774) per manufacturer's protocol. Protein-coupled beads were diluted and added to each well of Bio-Plex Pro 96-well Flat Bottom Plates (Bio-Rad). Beads were washed with PBS on a magnetic plate washer (MAG2x program) and 50uL of individual plasma samples diluted 1:300 in PBS were added to plate wells. A pool of pre-Covid-19 pandemic healthy human sera was used as a negative control (BioWest human serum AB males; VWR). Plates were incubated for 1 hour at room temperature with shaking, washed with PBS and incubated with 50uL of a 1:100 dilution of polyclonal Goat anti-human IgG-PE (OneLambda, Catalog LS-AB2) secondary antibody at room temperature for an additional 45 minutes with shaking. After incubation, samples were washed with PBS and resuspended in reading buffer and read on a Bio-Plex (Luminex) 200 plate reader with Bio-Plex Manager software (version 6.2; Bio-Rad) to obtain a mean fluorescence intensity (MFI) value for each sample. The MFI value for each plasma sample was divided by the mean value of the negative control samples to yield an MFI ratio. Based on negative control samples and samples from PCR-positive donors, seropositivity was determined based on MFI ratio cutoff values exceeding 6.0[19]. For this study, the lower limit of measured MFI ratios was restricted to 1, representing equivalent fluorescence intensity compared to negative control samples.

Luminex assay results were further validated by two commercial assays designed to detect anti-SARS-CoV-2 S- or N-specific total Ig (Elecsys Anti-SARS-CoV-2 S, Roche, Catalog

09289267190 and Elecsys Anti-SARS-CoV-2, Roche, Catalog 09203095190), respectively, using a Cobas e411 analyzer instrument (software version 03-02; Roche). Plasma samples from subsample participants were assayed according to manufacturer's instructions. In brief, 20uL of participant samples were incubated with a mixture of biotinylated and ruthenylated RBD antigen (for the S-assay) or a mixture of biotinylated and ruthenylated nucleocapsid antigen (for the N-assay) to form double-antigen-sandwich immune complexes. Afterward, streptavidin-coated magnetic microparticles were added. Within the measuring cell of the instrument, streptavidin microparticle-double-antigen-sandwich complexes were magnetically captured and washed. Voltage was applied to induce electrochemiluminescence (ECL) which was measured with a photomultiplier within the instrument, where increased ECL signals correspond to increased antibody titers. For the Roche Elecsys anti-S-Ig assay, values were expressed as U/mL. For the Roche Elecsys anti-N-Ig assay, values were expressed according to a cutoff index (COI). Seropositivity was determined based on cutoff values exceeding of a concentration of >0.8 U/ml for anti-S-Ig and a COI value of 1.0 for anti-N-Ig assays. An approximate conversion for anti-S-IgG MFI ratios obtained by Luminex assay to anti-S-Ig U/ml obtained using the Roche Elecsys Anti-SARS-CoV-2 S assay was determined using a linear regression model of log10-transformed data and can be found in[14].

Neutralization Assays

Cryopreserved plasma samples were thawed and evaluated for the presence of SARS-CoV-2 neutralizing antibodies using a cell- and virus-free assay[20] 50uL of diluted plasma samples (1:10, 1:30, 1:90, 1:270, 1:810, and 1:2430) were incubated with Luminex beads covalently coupled to the original SARS-CoV-2 Spike protein (2019nCoV) and Spike variants Delta (B.1.617.2) and Omicron (B.1.1.529) in Bio-Plex Pro 96-well Flat Bottom Plates (Bio-Rad) for 60 minutes at room temperature with shaking. Negative control wells on each plate included beads alone, and dilutions of pooled, pre-Covid-19 pandemic healthy human sera (BioWest human serum AB males; VWR). As a positive control, we included a high concentration (>1ug/mL) of two broadly neutralizing human monoclonal antibodies binding distinct epitopes on the SARS-CoV-2 S protein (Clones P2G3 and P5C3), isolated from previously infected and vaccinated donors[33,34]. After incubation, ACE2 mouse Fc fusion protein (produced by the Swiss Federal Institute of Technology in Lausanne (EPFL) Protein Production and Structure Core Facility) was added to each well at a final concentration of 1mg/uL and agitated for an additional 60 minutes. Beads were washed with PBS on a magnetic plate washer (MAG2x program) and 50uL polyclonal Goat F(ab') anti-mouse IgG-PE secondary antibody (Invitrogen, Catalog 12-4010-87) was added at a 1:100 dilution. Plates were incubated for 45 minutes at room temperature with shaking, washed, washed with PBS and resuspended in 80uL reading buffer and read on a Bio-Plex 200 plate reader with Bio-Plex Manager software (version 6.2; Bio-Rad). MFI values for beads alone without plasma or antibodies were averaged and used as the 100% binding signal for the ACE2 receptor to the bead-coupled spike trimer. MFI values from wells containing commercial anti-spike blocking antibodies were used as the maximum inhibition signal. Percent blocking of the spike protein trimer:ACE2 interaction was calculated using the formula: %inhibition=1-([MFI test dilution-MFI max inhibition]/[MFI max binding-MFI max inhibition])*100). A lower limit half maximal inhibitory concentration (IC50) serum dilution of 50 was set as the specificity cutoff using IC50 values of 104 pre-pandemic healthy donor samples (cutoff_50=12.5 mean IC_50+4*9.0 standard deviation (SD) to minimize detection of false-positive samples.

Interferon-gamma Release Assay (IGRA)

T cell responses were assessed by IGRA from stimulated whole blood. For the assay, 250uL of Li-heparin whole blood was plated per well in 96 well U-bottom plates (Sarstedt). Blood was stimulated for 20 hours in a humidified incubator at 37°C and 5% $CO_2$ with overlapping 15mer peptide pools spanning the entire M and N proteins (M/N pool) or the S1 domain of the spike protein and a mix of the predicted immunodominant peptides from the spike protein containing the majority of the S2 domain (S pool) (M, N, S1 and S PepTivator peptide pools, respectively; Miltenyi Biotec). Peptides were dissolved per manufacturer's instructions in sterile water and used at a final concentration of 0.6nmol (approximately 1ug/mLl) per individual peptide. As unstimulated negative controls, blood was incubated without peptide. As positive controls, blood samples were stimulated with 10mg/ml anti-CD3 antibody (Clone OKT3, Miltenyi Biotec, Catalog 130-093-387). After incubation, stimulated plasma was collected by 2x10min centrifugation at 500xg and stored at -20°C until analysis.

To evaluate the level of IFN-gamma in stimulated samples, the Human IFN-gamma ELISA assay (Human IFN-gamma DuoSet ELISA kit, R&D Systems, Catalog DY285B, and DuoSet ELISA Ancillary Reagent Kit 2, R&D Systems, Catalog DY008) was used according to manufacturer's instructions. In brief, assay plates were prepared by coating with 100uL of Capture Antibody (kit) overnight at room temperature. Plates were then washed, blocked with 300uL Reagent Diluent for 1 hours at room temperature, and then washed again. Cryopreserved, stimulated plasma was thawed and 100uL (or 100uL of IFN-gamma standard diluted serially 1:2 ranging from 600pg/mL to 9.4pg/mL) were added to each well of the prepared ELISA plates and incubated 2 hours at room temperature. Plates were then washed, 100uL of Detection Antibody was added and plates were incubated again for 2 hours at room temperature. Plates were again washed, then 100uL of Streptavidin-HRP B substrate were added for 20 minutes at room temperature, protected from light. Plates were washes, then 100uL of TMB Substrate was added. After 20 minutes incubated protected from light, 50uL of Stop Solution was added and plates were immediately read using a Multiskan SkyHigh Microplate Spectrophotometer with SkanIt software (Thermo Fisher Scientific) set at 450nm, with 540nm wavelength correction. Sample concentrations were interpreted from absorbances using a 4PL standard curve. Concentrations in unstimulated negative control wells were subtracted from the values of each test well and results were presented IFN-gamma pg/mL with negative values set to zero. Results were excluded if the background IFN-gamma concentrations in unstimulated control wells were greater than 5 times the median or if anti-CD3-stimulated positive control wells were negative.

The IGRA was compared to an ELISpot assay using the Human IFN-gamma ELISpot Assay kit (R&D Systems, Catalog EL285) following the manufacturer's instructions. For the assay, cryopreserved PBMCs were thawed in RPMI-1640 (Gibco, Thermo Fisher Scientific) and plated at 5e5 cells per well in in RPMI-1640 medium (Gibco, Thermo Fisher Scientific) supplemented with 5% human AB-serum (BioConcept) and 1% Penicillin-Streptomycin (Thermo Fisher) in assay plates. Cells were stimulated for 20 hours in a humidified incubator at 37°C and 5% $CO_2$ with M, N, S1 and S PepTivator overlapping 15-mer peptide pools at a final concentration of 0.6nmol (approximately 1μg/ml) per individual peptide, as for the IGRA (Miltenyi Biotec). As unstimulated negative controls, cells were incubated in culture medium alone, without peptide. As positive controls, 2.5e5 cells per well were stimulated with 10mg/ml anti-CD3 antibody (Clone OKT3, Miltenyi Biotec, Catalog 130-093-387). Spots were counted using an AID iSpot Reader System with EliSpot 7.0 software (AID). Two times the number of spots in unstimulated negative control wells were subtracted from the values of each test well and results were presented as spot-forming units (SFU) per 1e6 PBMCs with negative values set to zero. Results were excluded if background SFU counts in unstimulated control wells were greater than 5 times the median or if SFU counts in anti-CD3-stimulated positive control wells more 5 times less than the median.

Statistical Analyses

For the study population, for both demographic and self-reported SARS-CoV-2 vaccination coverage and previous positive Covid-19 tests, we reported percentages. For antibody and T cell positivity, we reported the percentage of the population which was positive. Being positive for antibody responses was considered as an MFI ratio above the detection cutoff (6.0 for S- and N-IgG and 50 for neutralizing activity half maximal inhibitory concentrations (IC50)). Being positive for T cell responses was considered as a value for pg/mL IFN-gamma produced greater than 0 after subtraction of background. We additionally reported geometric mean MFI ratio titers (S- and N-IgG), geometric mean IC50 values (neutralizing antibody responses to wildtype virus and delta and omicron variants) and geometric mean pg/mL IFN-gamma production (for S- and M/N-specific T cell responses) with corresponding 95% confidence intervals. Tests used for statistical comparisons between groups are indicated along with corresponding p values.

For assessing the correlation of antibody and T cell test results, we calculated Spearman correlation coefficients for all combinations of antibody subtypes and peptide-specific T cells, comparing the magnitude of responses as MFI ratios, IC50 values, or pg/mL IFN-gamma, respectively. Furthermore, we assessed concordance by calculating the proportion of participants testing positive or negative for antibody subtypes and overall T cells. Positive antibody responses were defined as an MFI ratio above the limit of detection (see above). Positive T cell responses were defined as a value for pg/mL of IFN-gamma >0 (see above). We also assessed agreement by calculating unweighted Cohen's Kappa values for test positivity for all possible combinations of antibody subtypes and virus-specific T cells.

Demographic and clinical factors associated with natural logarithm-transformed S- and N-IgG MFI ratio titers, neutralizing antibody IC50 values, and T cell IFN-gamma production were evaluated using multivariable linear regression models, adjusting for gender (female vs. male), age group (65+ vs. 16-64 years), reporting a previous positive Covid-19 test (yes vs. no), and the number of Covid-19 vaccine doses received (1, 2, 3+ vs. 0). To assess the relationship between S- and N-IgG titers, neutralizing antibody IC50 values, and T cell IFN-gamma production in March and reporting a positive Covid-19 test between March and June, a multivariable logistic regression model was constructed and adjusted Odds Ratios (OR) with 95% confidence intervals calculated. For each immune response variable, individuals were assigned to an expression quantile (i.e., expression for S-IgG was in the top 67% of all participants). Quantiles were calculated among all values >1 for each immune response variable and were rounded to the nearest value of 5. For S- and N-IgG MFI ratios, individuals were assigned to one of three expression quantiles (<33%, 33-67%, 67+% of all participants; S-IgG MFI ratios of Q1: 1 to <95, Q2: >=95 to <105, Q3: >=105; N-IgG MFI ratio titers of Q1: 1 to <5, Q2: >=5 to <10, Q3: >=10). Due to a larger range of expression values, for N-Ab IC50 values, and S- and M/N T cell IFN-gamma levels, individuals were assigned to one of four expression quantiles (<25%, 25->50%, 50->75%, 75+% of all participants; Wildtype-N-Ab IC50 values of Q1: 1 to <395, Q2: >=395 to <795, Q3: >=795 to <1535, Q4: >=1535; Delta-N-Ab IC50 values of Q1: 1 to <185, Q2: >=185 to <350, Q3: >=350 to <695, Q4: >=695; Omicron-N-Ab IC50 values of Q1: 1 to <100, Q2: >=100 to <175, Q3: >=175 to <360, Q4: >=360; S T cell IFN-gamma levels (pg/mL) of Q1: 1 to <25, Q2: >=25 to <65, Q3: >=65 to <155, Q4: >=155; M/N T cell IFN-gamma levels (pg/mL) of Q1: 1 to <10, Q2: >=10 to <30, Q3: >=30 to <95, Q4: >=95). Missing data were assumed to be missing completely at random and no imputation was performed.

To validate assay results, we compared Luminex S- and N-IgG results with anti-S and anti-N Ig assays using the Roche Elecsys system; we also compared S and M/N-specific T cell IFN-gamma responses obtained by IGRA with IFN-gamma ELISpot assay. For test comparisons, we calculated the percent concordance for categorical test results and Spearman correlation coefficients for absolute measurements. All study analyses were performed using Stata v.17.0 (StataCorp, LLC, College Station, TX, USA) and GraphPad Prism v.9.0 (GraphPad Software, Inc., San Diego, CA, USA). For all analyses, p values <0.05 were considered significant.

### SUPPLEMENTARY METHODS REFERENCES


1 Fenwick, C. *et al.* Changes in SARS-CoV-2 Spike versus Nucleoprotein Antibody Responses Impact the Estimates of Infections in Population-Based Seroprevalence Studies. *J Virol* **95** (2021). https://doi.org:10.1128/jvi.01828-20

2 Harris, P. A. *et al.* Research electronic data capture (REDCap)--a metadata-driven methodology and workflow process for providing translational research informatics support. *J Biomed Inform* **42**, 377-381 (2009). https://doi.org:10.1016/j.jbi.2008.08.010

3 Harris, P. A. *et al.* The REDCap consortium: Building an international community of software platform partners. *J Biomed Inform* **95**, 103208 (2019). https://doi.org:10.1016/j.jbi.2019.103208

4 Menges, D. *et al.* Heterogenous humoral and cellular immune responses with distinct trajectories post-SARS-CoV-2 infection in a population-based cohort. *Nature Communications* **13**, 4855 (2022). https://doi.org:10.1038/s41467-022-32573-w

5 Fenwick, C. *et al.* A high-throughput cell- and virus-free assay shows reduced neutralization of SARS-CoV-2 variants by COVID-19 convalescent plasma. *Science Translational Medicine* **13**, eabi8452 (2021). https://doi.org:doi:10.1126/scitranslmed.abi8452

6 Fenwick, C. *et al.* A highly potent antibody effective against SARS-CoV-2 variants of concern. *Cell Reports* **37** (2021). https://doi.org:10.1016/j.celrep.2021.109814

7 Fenwick, C. *et al.* Patient-derived monoclonal antibody neutralizes SARS-CoV-2 Omicron variants and confers full protection in monkeys. *Nature Microbiology* **7**, 1376-1389 (2022). https://doi.org:10.1038/s41564-022-01198-6
