## Supplementary Figures for "Longitudinal Humoral and Cell-Mediated Immune Responses in a Population-Based Cohort in Zurich, Switzerland between March and June 2022 - Evidence for Protection against Omicron SARS-CoV-2 Infection by Neutralizing Antibodies and Spike-specific T cell responses"

**Supplementary Figure 1. Flow Chart of Study Participation and Testing.** A total of 1044 individuals participated in our March 2022 analysis. All were tested for antibody responses. 328 participants were also tested for T cell responses. Of the initial 1044 participants enrolled, 964 returned for a follow-up analysis in June 2022. All were tested for antibody responses. 141 participants were also tested for T cell responses. Of the 141 tested for T cell responses in June 2022, 118 had been also tested in March 2022 and 23 had not been previously tested.

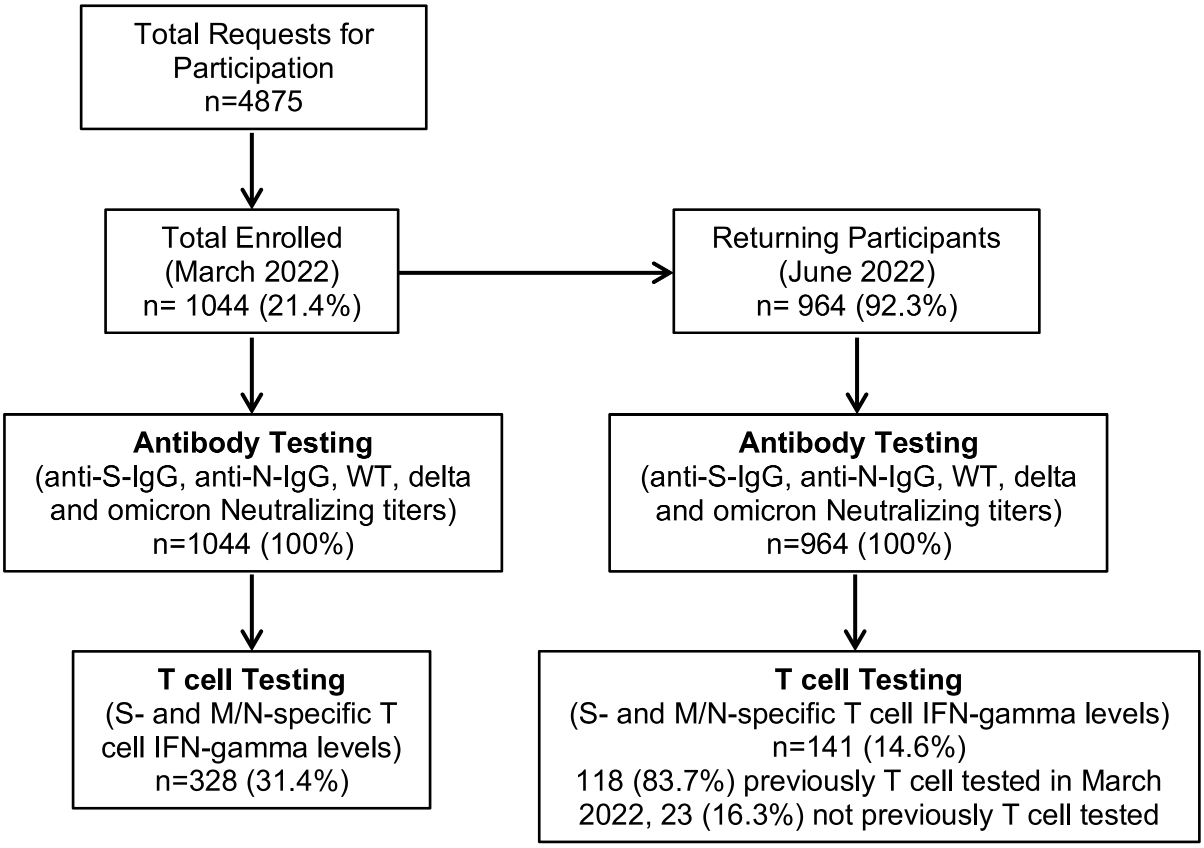

**Supplementary Figure 2. Concordance between Antibody and T cell Responses among Participants.** A) Percentage of participants with complete data, seropositive for anti-S- and -N-IgG titers and S- and M/N-T cells in March 2022 (n=328). B) Percentage of participants with complete data, seropositive for anti-S- and -N-IgG titers and S- and M/N-T cells in June 2022 (n=141).

**A**

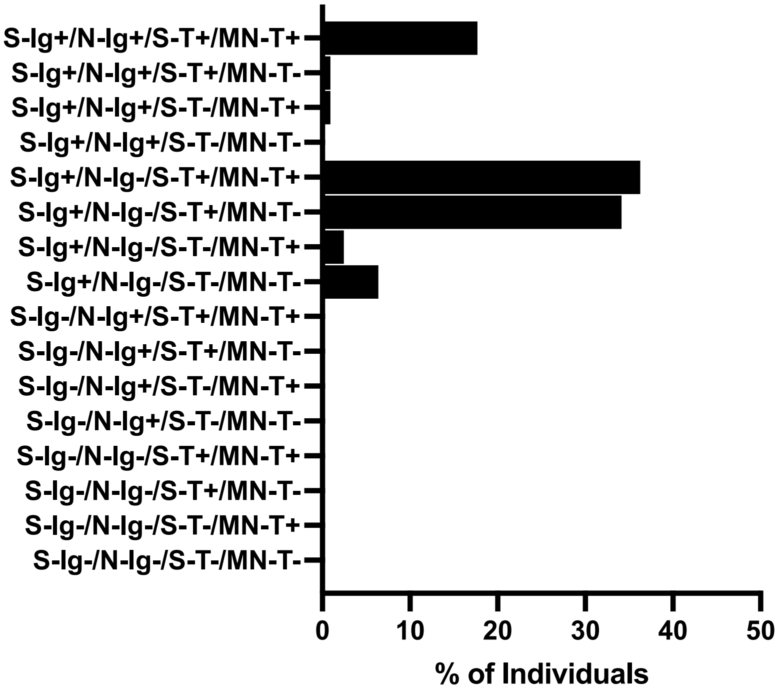


**B**

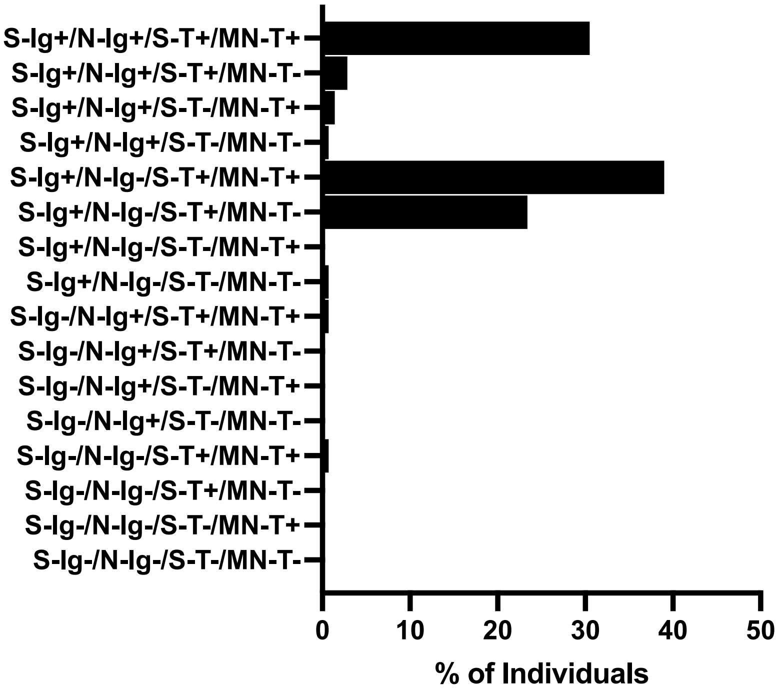

**Supplementary Figure 3. Agreement and Correlation between Luminex and Roche Antibody Assays and IGRA and ELISpot T cell Assays.** A) Percentage agreement between results of listed assay types (n=50 for antibody assays, n=48 for T cell assays). B) Spearman correlation coefficients for comparisons between listed assay types (n=50 for antibody assays, n=48 for T cell assays).

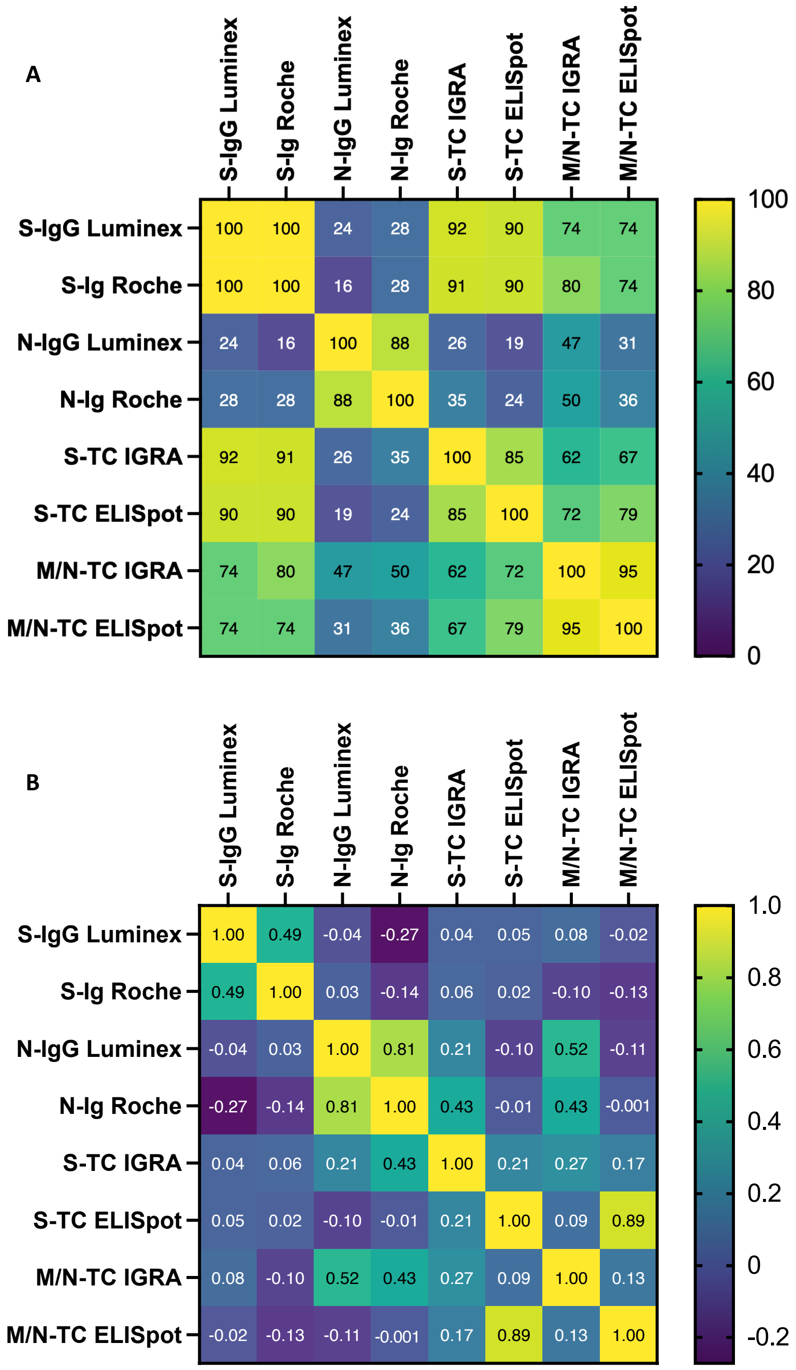
