## Supplementary Table for "Longitudinal Humoral and Cell-Mediated Immune Responses in a Population-Based Cohort in Zurich, Switzerland between March and June 2022 - Evidence for Protection against Omicron SARS-CoV-2 Infection by Neutralizing Antibodies and Spike-specific T cell responses"

**Supplementary Table 1. Participant Demographics**

| | March 2022 (n=1044) | | June 2022 (n=964) | |
|---|---|---|---|---|
| **Sex** | **n** | **%** | **n** | **%** |
| Male | 475 | 45.6 | 438 | 45.5 |
| Female | 567 | 54.4 | 524 | 54.5 |
| Other/Unknown | 2 | - | 2 | - |
| **Age** | **n** | **%** | **n** | **%** |
| 16-64 | 769 | 73.6 | 697 | 72.3 |
| 65+ | 275 | 26.3 | 267 | 27.7 |
| Unknown | 0 | - | 0 | - |
| **Vaccine Doses** | **n** | **%** | **n** | **%** |
| 0 | 68 | 6.6 | 62 | 6.4 |
| 1 | 28 | 2.7 | 23 | 2.4 |
| 2 | 194 | 18.7 | 165 | 17.1 |
| 3+ | 750 | 72.0 | 714 | 74.1 |
| Unknown | 4 | - | - | - |
| **Positive Test (Pandemic Start - March 2022)** | **n** | **%** | **n** | **%** |
| Yes | 339 | 32.6 | 154 | 16.0 |
| No | 702 | 67.4 | 810 | 84.0 |
| Unknown | 3 | - | - | - |
| **Positive Test (Pandemic Start - June 2022)** | **n** | **%** | **n** | **%** |
| Yes (First Infection) | - | - | 132 | 13.7 |
| Yes (Repeat Infection) | - | - | 306 | 31.8 |
| No | - | - | 525 | 54.5 |
| Unknown | - | - | 1 | |
